# Detecting known neoepitopes, gene fusions, transposable elements, and circular RNAs in cell-free RNA

**DOI:** 10.1101/2024.06.07.24308622

**Authors:** Mayank Mahajan, Martin Hemberg

## Abstract

Cancer is the second leading cause of death worldwide, and although there have been advances in treatments, including immunotherapies, these often require biopsies which can be costly and invasive to obtain. Due to lack of pre-emptive cancer detection methods, many cases of cancer are detected at a late stage when the definitive symptoms appear. Plasma samples are relatively easy to obtain, and they can be used to monitor the molecular signatures of ongoing processes in the body. Profiling cell-free DNA is a popular method for monitoring cancer, but only a few studies have explored the use of cell-free RNA (cfRNA), which shows the recent footprint of systemic transcription. Here we developed FastNeo, a computational method for detecting known neoepitopes in human cfRNA. We show that neoepitopes and other biomarkers detected in cfRNA can discern Hepatocellular carcinoma (HCC) patients from the healthy patients with a sensitivity of 0.84 and a specificity of 0.79. For colorectal cancer we achieve a sensitivity of 0.87 and a specificity of 0.8. An important advantage of our cfRNA based approach is that it also reports putative neoepitopes which are important for therapeutic purposes.

## INTRODUCTION

Cell-free nucleic acids are either released by dying cells or actively secreted by live cells into body fluids, such as plasma. These nucleic acids can be acquired via liquid biopsy and sequenced to study the active processes in the body, and by detecting mutations or epigenetic modifications one can infer the presence of a tumour (1, 2). Consequently, cell-free DNA (cfDNA) is emerging as a tool for disease monitoring, and detection of early-stage tumours (3, 4). By contrast, cell-free RNA (cfRNA) is less well studied even though it can reveal actively transcribed of genes, mutations, repetitive elements, and exosomal RNA from the whole body (5–9). cfRNA shows a current footprint of the systemic RNA expression, however the tissue of origin of the cfRNA cannot be ascertained easily. It has been estimated that up to 95% of the cfRNA in plasma is derived from other tissues or the microbiome, with only a small minority originating from the tumour (10). Thus, deriving clinically relevant information from cell-free RNA is challenging.

We describe an approach which considers point mutations and gene fusions to detect putative neoepitopes in cfRNA. Epitopes are short peptide fragments derived from proteins that are expressed in a cell. Some of these epitopes bind to the human leukocyte antigen (HLA) class I or class II molecules and are then presented on the cell surface for immune surveillance. Mutations in the coding sequence can give rise to neoepitopes that are missing in the proteome of healthy germline cells. Cancer-specific neoepitopes are ideal targets for immunotherapies since they are unique to malignant cells, and they are hence highly sought after (11, 12).

Gene fusions have been shown to be recurrent in some cancer types and they can be used as diagnostic biomarkers (13, 14). Gene fusions can produce neoepitopes which have been explored in various studies (15–17). Importantly, T cell response against these gene-fusion neoepitopes can be used for personalised immunotherapies to target cancers that are driven by gene fusions (18, 19).

Retrotransposons are typically silenced in mammalian cells, nonetheless 6-30% of mammalian transcripts have been shown to initiate within repetitive elements (20). Disruption of the epigenetic landscape in cancer can result in upregulation, and expression of long interspersed nuclear elements (LINE) and human endogenous retrovirus (HERV) have been observed in various cancer types (21, 22).

Circular RNAs (circRNAs) are regarded as reliable biomarkers for cancer (23–25). circRNAs are resistant to exoribonuclease (RNase R) and their closed structure provides higher stability compared to their linear counterparts (24). circRNAs have been detected in circulating exosomes to show their potential as biomarkers for cancer diagnosis (26, 27)(Li et al., 2015; Roy et al., 2022).

Here we quantify the expression of neoepitopes, gene fusions, transposable elements (TEs), and circRNAs in the cfRNA and demonstrate that together these features can be used to distinguish cancer patients from healthy controls with high sensitivity and specificity.

## MATERIAL AND METHODS

### Nullomers associated to neoepitope databases

Neoepitopes described in Immune epitope database (IEDB) and Tumor-Specific NeoAntigen database (TSNAdb) were acquired (28, 29). The ‘epitope full v3.csv’ file was downloaded from IEDB webpage www.iedb.org, on Sept-19, 2022. Neoepitopes and wild-types epitopes with the following characteristics were retrieved from the above file: ‘RelatedObject EpitopeRelationship’ is ‘neoepitope’, ‘RelatedObject OrganismName’ contains the word ‘sapien’, ‘Epitope Description’ only has letters A to Z, ‘RelatedObject ParentProteinIRI’ has the uniprot ID. The uniprot IDs in IEDB were converted to transcript IDs using ‘HUMAN 9606 idmapping selected.tab’, which was downloaded from www.uniprot.org on Jun-30, 2022. 114 uniprot IDs in IEDB did not have a corresponding transcript ID. Next, ‘frequent_neoantigen_ICGC_4.0_adj.txt’ and ‘frequent_neoantigen_TCGA_4.0_adj.txt’ were downloaded from TSNAdb webpage http://biopharm.zju.edu.cn on Sept-19-2022, and the neoepitopes were retrieved along with the corresponding wildtype epitopes and transcript IDs.

To find the position of neoepitopes on the human consensus coding sequence (CCDS), each wild-type epitope was searched on the protein translation of the corresponding CCDS. The neoepitopes were then aligned to the wild-type epitopes using the Smith-Waterman local alignment algorithm and the aligned sequences were used to detect the sites of insertion, deletion, and substitution. 521 unique neoepitopes from IEDB that had an insertion or deletion of amino acid, or >2 amino acid substitutions were excluded (Supplementary Table S1). The TSNAdb neoepitopes consist of single nucleotide substitutions only.

Possible DNA conformations corresponding to each neoepitope were generated using all possible combinations of codons at the sites of mutation. The sequence around the mutated sites in each DNA conformation was scanned for CCDS-nullomers of length 16, which are nucleotide kmers of length 16 that are not present in the human CCDS. The length of the nullomers was chosen to provide a balance between sensitivity and specificity of the detection algorithm and to provide faster search speed (3). The CCDS-nullomers could be present in other parts of the genome and result in false positive matches from other transcribed parts of the genome, such as repeat elements. To improve the specificity to CCDS, nullomers that appear >300 times in the human genome sequence were removed. The above filter eliminates few neoepitopes, as 99.9 % of the neoepitope-derived nullomers occur <139 times in the human genome sequence (Supplementary Figure S1). Unless otherwise mentioned, Ensembl GRCh38.p13 genome release 107 was used for human genome sequence, CCDS and proteins.

### Neoepitope binding affinity to HLA complexes

netMHCpan and netMHCIIpan were used to predict the binding affinity of both neoepitopes and wildtype epitopes to various MHC complexes (30, 31). Since TSNAdb epitopes are 8-11 amino acids (aa) long, netMHCpan was used to predict the binding affinity to MHC I complexes. We used the default list of MHC I alleles in the TSNAdb and added 3 more alleles, HLA-C*08:01, HLA-A*33:03, HLA-B*08:01, which have high population frequencies as per the Allele Frequency Net Database (28, 32).

IEDB has both long and short neoepitopes. The binding affinity of neoepitopes of length <= 11 aa was predicted against the MHC I complex using the same approach that was used for the TSNAdb neoepitopes. In addition, netMHCIIpan was used to predict the binding affinity of neoepitopes of length > 11 aa to the MHC II complex. We predicted their binding affinity against the following MHC II alleles that were chosen for high frequency:-DRB1*0701, DRB1*1501, DRB1*0301, DRB1*1101, DRB1*0101, DRB1*1302, DRB1*1301, DRB1*1502, DRB1*0401, DRB1*1201, DRB1*0403, DRB4*0101, DRB3*0202, DRB3*0101, DRB5*0101, DRB3*0301, HLA-DQA1*0102-DQB1*0501, HLA-DQA1*0103-DQB1*0501, HLA-DQA1*0501-DQB1*0201, HLA-DPA1*0103-DPB1*0201, HLA-DPA1*0103-DPB1*0402 (32–36).

Only HLA binding predictions with BindLevel equal to ‘SB’ or ‘WB’ were retained and the HLA alleles and binding affinities were added to the respective neo- or wildtype epitopes in the mapping file described above. In case of multiple predictions per epitope, all the predictions are shown.

### Neoepitope frequency in healthy population

The germline variants form various genetic ancestry groups, as described using exome sequencing in gnomAD v4.0.0, were used to extract population frequency of each neoepitope producing mutation (37). Allele frequency in females (AF_XX), males (AF_XY), Africans/African Americans (AF_afr), admixed Americans (AF_amr), Ashkenazi jews (AF_asj), east Asians (AF_eas), Finnish (AF_fin), middle eastern (AF_mid), non-finnish europeans (AF_nfe), south asians (AF_sas), combined allele frequency (AF), and the genetic ancestry group with highest proportion of the allele (grpmax), were retrieved from the gnomAD database for the allelic variants using the following criteria: AF >1e-07, VEP:Consequence = ‘missense_variant’, VEP:BIOTYPE = ‘protein_coding’, VEP:Feature consists of a transcript ID starting with ‘ENST’, and the allelic variant produces one of neoepitopes described in the above section. The population variants data retrieved from the gnomAD database was then added to the respective codon sequences of neoepitopes in the mapping file described above.

### Nullomers associated to gene fusions

Gene fusion file “ChimerKB4.xlsx” was downloaded from ChimerDB and selected a subset of gene fusions with ‘ChimerPub’ column = ‘Pub’ (May-19-2023) (38). 500 nucleotides were extracted from upstream of the 3’ end and downstream of the 5’ end of the fusion junction and we fused them at the junction. The nucleotide sequences were extracted from the same human genome sequence that has been used to generate the ChimerDB (downloaded from ‘hgdownload.cse.ucsc.edu/goldenpath/hg19/bigZips/’). The fused sequences generated here are used to map the reads in the cfRNA search pipeline. We searched CCDS-nullomers that spanned the fusion junctions, and removed the ones that were repeated > 300 times in the genome. We scanned neoepitopes at the fusion junctions where 1) both 5’ and 3’ junctions are inside the coding sequence, 2) the 5’ junction does not overlap the start codon, and 3) the 3’ gene is still in its original frame after fusion. 9 aa long peptides that were absent from the reference human protein sequences were classified as neoepitopes. We ignored neoepitopes produced via fusions in the intron and UTRs regions as it is non-trivial to predict the neoepitopes produced by such fusions.

### Neoepitope and fusion detection pipeline

The pipeline uses stranded RNA-seq data from cfRNA and CCDS-nullomers generated in the above section as input and outputs the putative neoepitopes and gene fusions detected in the cfRNA (Figure 1).

**Figure 1.**
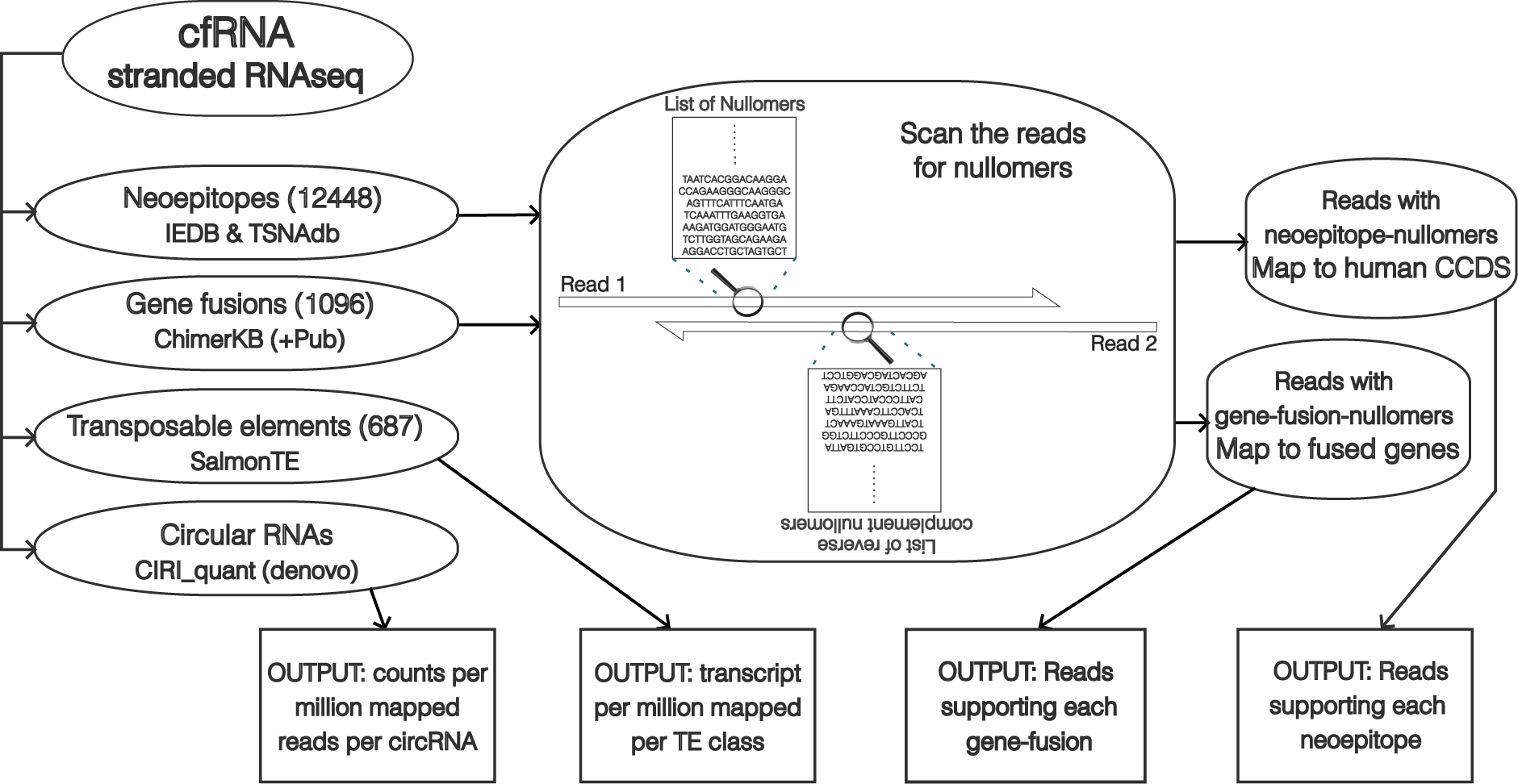
Overview of the FastNeo pipeline.

Step 1: The nullomers of length 16 are searched on the first read, and reverse complement sequences of those nullomers were searched on the second read of the paired-end RNA-seq sequencing data.

Step 2: Read pairs containing nullomer/nullomers associated to neoepitope databases are mapped to the coding sequences of the human genome GRch38 using bowtie2 with parameters (–very-sensitive-local) (39). Duplicate reads are then removed using samtools rmdup (40). Additionally, reads with more than 33.3% soft-clipped bases are removed. All reads with MAPQ score >10, or the alignment score (AS) greater than a minimum expected score (MES) are retained.

MES is a linear function of aligned length (AL) defined as, MES = slope * (AL - 35) + 64, where slope is calculated using the desired AS when the AL is between 35 and 150 nucleotides. We used AS =64 for AL =35, which allows 1 medium quality mismatch (penalty=4) and AS =277 for AL =150 ∼4 medium quality mismatches (penalty=15); hence, slope = (277–64) / (150–35) = 1.85.

Reads pairs containing >=1 nullomers associated with gene fusions are mapped to the fused sequences created above using bowtie2 with parameters (– very-sensitive-local) (39). Duplicate reads are then removed using samtools rmdup (40). Additionally, reads with more than 33.3% soft-clipped bases are removed and only reads with 5 or more bases mapped on both sides of the fusion junction are retained. MAPQ scores are less relevant when reads are mapped to a very small set of sequences. Hence, only reads with MAPQ score >30 or AS greater than the MES, as described above, are retained.

Step 3: The read coverage per nullomer is calculated as the total reads that contain the nullomer sequence and are mapped to the corresponding neoepitope coding sequence or gene fusion junction. The read coverage of each neoepitope and gene fusion is calculated as the read coverage of the most covered corresponding nullomer. The total nullomers found per neoepitope or gene fusion are also reported. The neoepitopes produced by the same mutation are reported together in the same row.

*Output.* The neoepitope output file shows 1) the fastq file name, 2) gene id 3) HGNC symbol, 4) most covered nullomer per mutation/mutations, 5) neoepitopes associated to the nullomer, 6) number of mapped reads, 7) number of nullomers on the read with most nullomers, 8) database name, 9) gene function, HLA binding affinities of wildtype epitope, 10) and neoepitope, and 11) the frequency of the neoepitope producing mutations in healthy individuals of various genetic ancestry groups.

The gene fusion output file shows 1) fastq file name, 2) ChimerKB ID, 3) most covered nullomer per gene fusion, 4) neoepitopes associated with the gene fusion, 5) number of mapped reads, 6) number of nullomers on the read with most nullomers, 7) gene function, 8) genomic loci of 5’ junction, and 9) genomic loci of 3’ junction.

The read coverage of variants is low in the cfRNA sequencing data, hence low threshold of filters was used. Neoepitopes with >=3 mapped reads were used for all the downstream analyses. Fusions with >=2 mapped reads and >=2 detected nullomers were used for all the downstream analyses.

### Benchmarking neoepitope detection against other variant calling methods

Known neoepitopes were detected in the cfRNA samples in GSE142987 and GSE136651 datasets using GATK HaplotypeCaller (41), samtools/bcftools mpileup (42), and Lofreq (43), which specifically suited to low coverage datasets (44). We mapped all the reads to the human genome using STAR 2.7.10a (45) with the following options: -outSAMmultNmax 3 - outFilterMultimapNmax 15 -outFilterMismatchNmax 15 -alignSJDBoverhangMin 2 - alignIntronMax 1000000 -peOverlapNbasesMin 10 Values used for outFilterMultimapNmax, alignIntronMax and alignSJDBoverhangMin were inspired from the section ‘ENCODE options’ in the STAR manual. Option peOverlapNbasesMin is said to improve mapping accuracy for paired-end libraries with short insert sizes. GATK ‘MarkDuplicates’ tool was used to remove the duplicate reads (http://broadinstitute.github.io/picard/).

For bcftools, the mapped reads were passed to ‘bcftools mpileup’ with options ’-q 10 -Q 30’, and ‘bcftools call’ was used to call variants. The variants supported by <3 reads were filtered out. For Lofreq and HaplotypeCaller, base qualities of mapped reads were re-estimated by running ‘SplitNCigarReads’, ‘BaseRecalibrator’ with option ’-known-sites 1000GENOMES- phase_3.vcf’, and ‘ApplyBQSR’ in that order. These tools are part of the GATK and ’1000GENOMES-phase_3.vcf’ file containing allele frequencies from 1000 genomes phase 3 populations was downloaded from the ensembl ftp site. The mapped reads with recalculated base quality were then passed to ‘lofreq call’ and ‘HaplotypeCaller’ with option ’--dbsnp ’1000GENOMES-phase_3.vcf’ for calling variants. For HaplotypeCaller, variants with FS > 30.0, QD < 3.0 and AD < 3 were filtered out. Ensembl VEP (46) was used to find missense and frameshift variations among the variants discovered by HaplotypeCaller, bcftools and Lofreq. The neoepitopes in IEDB/TSNAdb were detected around the missense and frameshift mutations using custom scripts (available in the FastNeo github page).

The run-time of all the tools were compared on the cfRNA from GSE142987 dataset, with average 8 million bases per sample, and GSE136651 dataset, with average 0.7 million bases per sample. Each sample was run on 2 cores making use of both cores in the steps that permitted use of multiple cores.

To use STAR instead of bowtie2 in FastNeo, the reads with detected nullomers in the cfRNA samples in GSE142987 and GSE136651 datasets were mapped to the human genome using STAR 2.7.10a (45) with same options as above. Duplicate reads were removed using GATK ‘MarkDuplicates’ tool (http://broadinstitute.github.io/picard/). The sam files with the remaining mapped reads were transformed from genome coordinates to the CCDS coordinates using ‘mudskipper shuffle’ and ‘mudskipper bulk -p’ (https://github.com/OceanGenomics/mudskipper).

### Data Sets

We used four plasma-derived cfRNA datasets (Supplementary Table S1). The first consists of 30 healthy donors and 35 HCC patients (GEO id: GSE142987) (6). The second dataset consists of 20 healthy donors, 8 HCC patients and 10 multiple myeloma patients (GEO id: GSE182824) (47). The second dataset also includes samples from pre-cancerous conditions that were ignored in this study. The third dataset consists of 46 healthy donors and 54 colorectal, 37 stomach, 27 HCC, 35 lung, and 31 esophageal cancer patients (GEO id: GSE174302) (8). The fourth dataset consists of 11 of pancreatic cancer patients and 22 healthy donors (GEO id: GSE136651) (7). All data was de-identified, and there is no metadata about the treatment these patients were undergoing.

All datasets were sequenced using stranded RNA-seq libraries. Data from GSE142987, GSE182824 and GSE174302 has been sequenced using Illumina HiSeq with similar throughputs of ∼9 million bases, while data from GSE136651 has been sequenced using Illumina NextSeq with a much lower throughput of ∼0.7 million bases (Supplementary Table S1). Hence the data from GSE136651 was analysed separately, while the other three were analysed together.

### Discriminative neoepitopes

FastNeo was used to quantify neoepitope expression in cfRNA datasets. The read coverage of each neoepitope was normalised by the total reads in the corresponding sample and then scaled by log10 to estimate the mean normalised expression per neoepitope. Neoepitopes that were present >20% of all the cancer patients, >20% healthy donors in the merged dataset, and had a mean normalised expression >0.1 in both groups of donors were categorised as generic neoepitopes. Generic neoepitopes were removed from all the analyses and figures (Supplementary Table S2). Neoepitopes with twice the mean expression in cancer patients as compared to the healthy donors, or vice-versa were categorised as discriminative neoepitopes. In addition, discriminative neoepitope should be detected in cfRNA of >=3 individuals from the group they are enriched in. For ease of visualization, the discriminative neoepitopes that are shown in the Figure 2 and S3 were detected in cfRNA of >=5 individuals from the group they are enriched in. The heatmaps of discriminative neoepitopes were plotted using pheatmap library in R and the neoepitopes were clustered using ‘ward.D2’ method and the euclidean distance.

**Figure 2.**
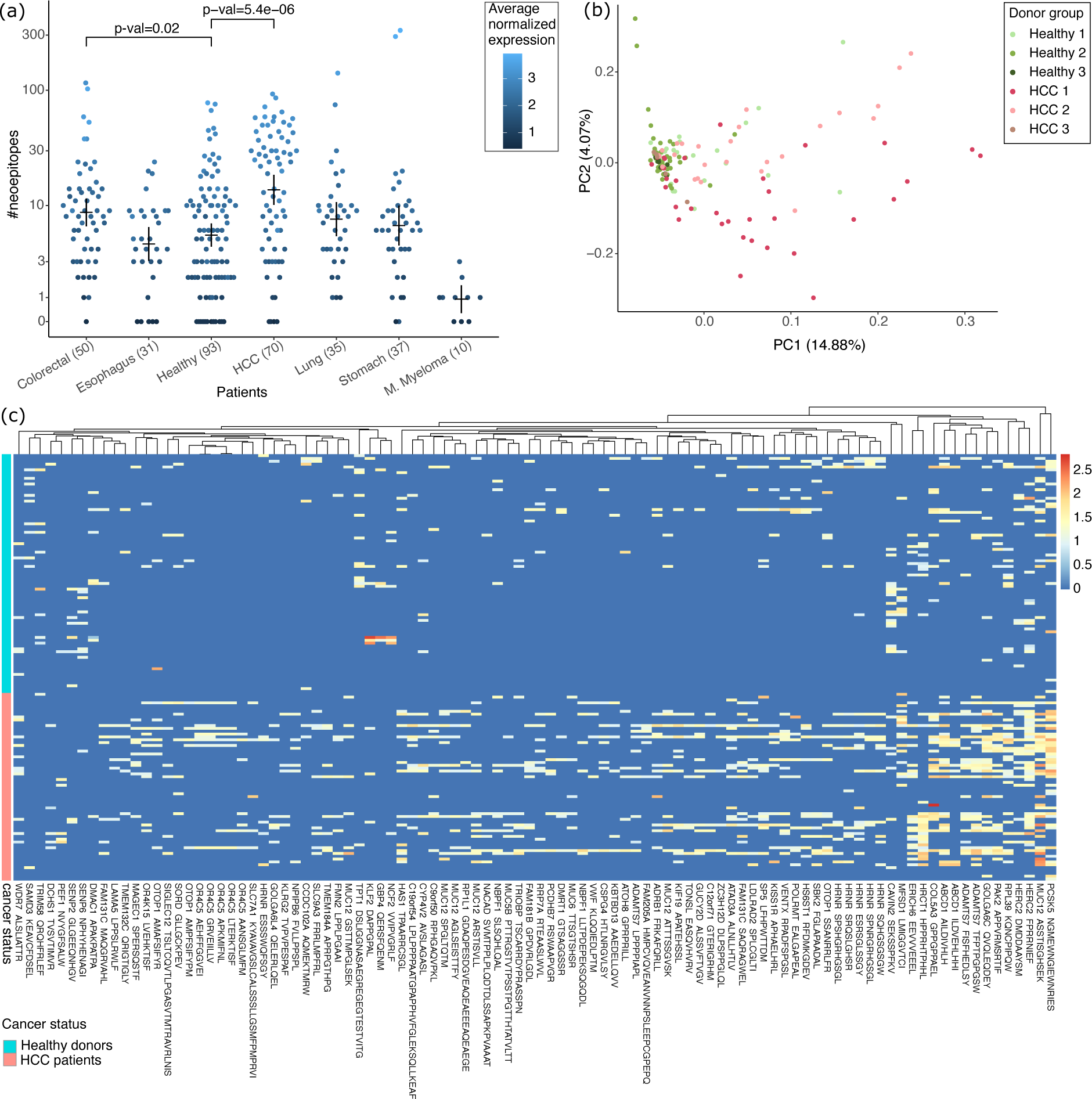
Putative neoepitopes detected in cfRNA. (a) Number of known neoepitopes detected in plasma cfRNA of patients grouped by cancer type and healthy. Each dot is coloured by average normalized expression for all the putative neoepitopes in the respective donor. P-values show the significance of the difference in distribution of healthy donors vs. patients from two cancer types with most donors, calculated using the Wilcoxon test. Means and error-bars are shown for each group. (b) First two components of a PCA using expression of all putative neoepitopes in HCC patients and healthy donors. Suffixes 1, 2 and 3 in the donor groups correspond to the different datasets in Supplementary Table S1. (c) Heatmap showing the expression of discriminative neoepitopes detected in the cfRNA of cancer patients and healthy donors (see Methods). Each column represents a neoepitope, and each row represents a patient or a healthy donor. Column label shows one of the neoepitopes together with the gene symbol. See Supplementary Table S5 for a full list of discriminative neoepitopes with descriptions.

### Discriminative TEs

SalmonTE (parameters: quant --reference=hs) was used to quantify transposable elements in cfRNA datasets (48). SalmonTE orders human TEs into 687 manually curated classes, and the expression is measured for each class of TE. SalmonTE output consists of effective length, reads mapped, and transcript per million (TPM) for each of the 687 TE classes (Figure 1). Reads per kilobase per million mapped reads (RPKM) values were calculated to measure the expression of each TE class in each sample using the effective length, reads mapped and the total bases in the sample.

The pairwise Wilcoxon test and Bonferroni correction was performed using log10 of RKPM of each TE class to detect differentially expressed TEs in the specific cancer type vs. healthy donors. TEs were highly expressed in cfRNA samples, hence the TEs expressed in <80% of healthy donors or <80% of cancer patients were removed before performing the test. The p-value based cut-offs used to select most discriminative TEs.

### Discriminative circRNAs

CIRIquant 1.1.3 was installed with default settings and gencode hg19 genome as specified in the CIRIquant documentation (15), and was run with parameter ‘--library-type 1’ to quantify the circRNA in cfRNA datasets. CIRIquant reports circRNA expression in count per million (CPM), which is already normalised against overall RNA expression. circRNAs with <2 reads mapped to the back splicing junction were removed from all downstream analysis.

The pairwise Wilcoxon test and Bonferroni correction was performed using log10 of CPM scores of each gene to detect differentially expressed circRNA genes in the specific cancer type vs. healthy donors. circRNA genes expressed in <40% of the samples were removed before performing the test. In case of multiple circRNAs per gene, the circRNA with highest CPM score was used. The p-value based cut-offs used to select most discriminative circRNAs.

### Classification of cancer patients

The supervised learning models were trained on normalised and log-scaled neoepitope expression to classify cancer patients against the healthy individuals. The generic neoepitopes, as described above, were ignored for training classifiers. The feature selection was done separately for each fold. The selected neoepitopes must have twice the mean expression in cancer patients as compared to the healthy donors, or vice-versa, and must be detected in cfRNA of >=3 individuals of the group they are enriched in. Five most discriminative TEs with lowest p-values, and p-value < 1e-7 were selected using pairwise Wilcoxon test and Bonferroni correction. TEs expressed in less than 80% of healthy donors or <80% of patients of the specific cancer type were removed before performing the pairwise Wilcoxon test. Five circRNAs with lowest p-values, and p-value < 1e-4 were selected using pairwise Wilcoxon test and Bonferroni correction. circRNAs expressed in less than 40% of healthy donors or patients of the specific cancer type were removed before performing the pairwise Wilcoxon test.

The classifiers were trained in 10 independent randomised iterations using 5-fold cross validation on general linear models with Ridge and Lasso regularizations, support vector machine (SVM) and Random Forests. The performance metrics achieved by each method was averaged across all folds first and then across all iterations. The R library ‘glmnet’ was used to train linear regression models using Ridge and Lasso regularizations with options family=’binomial’. Lambda values were optimised separately for Ridge and Lasso using ‘cv.glmnet’. All values of lambda between 10^2^ and 10^-2^ with step change of 10^-0.1^ were tested. Eventually, lambda=0.6 for Ridge and lambda=0.04 for Lasso was used for classification using only neoepitopes, and lambda=0.1 for Ridge and lambda=0.03 for Lasso was used for classification using other feature sets. R library ‘randomForest’ was used to train random forest models with option nodesize = 2. R library ‘E1071’ was used to train the SVM models with options kernel = linear, cost=2 and scale = FALSE. The cost=2 was used for SVM to prevent overfitting.

## RESULTS

### Neoepitope and gene-fusion associated nullomers

To increase the likelihood of identifying bona fide neoepitopes we restrict our search to the ones described in the TSNAdb and IEDB (28, 49). TSNAdb consists of neoepitopes that are present in four or more cancer patients in the The Cancer Genome Atlas (TCGA) and International Cancer Genome Consortium (ICGC) databases. IEDB consists of neoepitopes that have been experimentally characterised in published studies and many of them have also been tested for immune response using methods such as qualitative binding to HLA complexes and assays to monitor the release of IFNg, TNFa, and various interleukins. The experimental data associated with IEDB neoepitopes can be explored using the interactive platform CEDAR (50).

To identify putative neoepitopes and gene fusions from cfRNA, we utilise nullomers, kmers that are not present in the human reference sequence. Here, we employ the CCDS-nullomers, kmers that are absent in the consensus coding sequence (CCDS) (51). As CCDS-nullomers are not expected in the consensus coding sequence, they can be deployed for a fast, yet reliable search of neoepitope producing mutations and cancer detection (3). Most neoepitopes and gene-fusions result in one or more CCDS-nullomers being created, and thus, they are well suited for nullomer-based detection.

We scanned the neoepitopes from TSNAdb and IEDB for CCDS-nullomers of length 16. As these neoepitopes have been described using different versions of the genome, the transcript ID of some genes could not be found in the CCDS fasta, and some of the wildtype epitopes could not be found in the corresponding protein coding sequence in GRCh38.p13 (Supplementary Table S3a). We recovered 12,526 out of 13,582 (∼92%), and 79,871 out of 85,185 (∼94%) unique combinations of wildtype epitopes and neoepitopes from IEDB and TSNAdb, respectively (Supplementary Table S3a). In addition, <1% of the neoepitopes did not contain any CCDS-nullomers of length 16 in their coding sequence (Supplementary Table S3a). To reduce false positives, we characterised the neoepitopes associated with germline variants from the genetic ancestry groups described in the gnomAD database (37, 52). 645 and 9,811 neoepitopes in IEDB and TSNAdb, respectively, were associated with germline variants, of which the african/african american (AFR), non-finnish european (NFE) and south asian (SAS) were three most frequent ancestry groups (Supplementary Table S4a).

IEDB had 3,661 unique neoepitopes of length <= 11 aa and 8,787 unique neoepitopes of length 12-37 aa (Supplementary Table S3b). The binding affinity of the shorter neoepitopes was predicted against the MHC I complex (see Methods), while binding affinity of the longer neoepitopes was predicted against the MHC II complex (Supplementary Table S3b). 2,833 out of 3,661 neoepitopes in IEDB and 69,095 out of 79,871 TSNAdb were predicted to have binding affinity to one or more MHC I alleles. HLA-A*02:01, HLA-C*01:02 and HLA-C*04:01 were predicted as the three most frequent HLA alleles that bind to IEDB neoepitopes, while HLA-C*07:01, HLA-C*07:02 and HLA-C*04:01 were predicted as the three most frequent HLA alleles that bind to TSNAdb neoepitopes (Supplementary Table S4b). Alas, only 13 out of 8,787 neoepitopes in IEDB were predicted to have binding affinity to one or more MHC II alleles. This was not surprising since MHC II binding prediction is still an emerging field and has not been studied as much as MHC I binding.

Among the gene fusions in the ChimerKB dataset that are supported by published literature, we were able to retrieve 1,096 unique gene fusion junctions. Nine or more CCDS-nullomers of length 16 were retrieved from each of the 1,096 gene fusion junctions, and 182 of these junctions were found to generate putative neoepitopes.

### Neoepitopes in cfRNA

Nullomers were used to detect putative neoepitopes in the plasma cfRNA of cancer patients and healthy donors in five public datasets that were sequenced using stranded RNA-seq libraries and contained >=10 samples per condition. cfRNA for most cancer types in this study is derived from GSE174302, except multiple myeloma, which is derived from GSE182824, and hepatocellular carcinoma (HCC), which is merged from three different datasets (Supplementary Table S1). The healthy donors were also merged from three different datasets (Supplementary Table S1).

A median of only one putative neoepitope was discovered in cfRNA of multiple myeloma patients. By contrast, a median of 21 putative neoepitopes were discovered in cfRNA of HCC patients (Figure 2a). This difference was not surprising as tumour mutational burden is generally higher in HCC patients (53, 54). Surprisingly, a median of 6 putative neoepitopes were discovered in the cfRNA of healthy donors. Although the number of putative neoepitopes discovered in HCC patients was noticeably higher than in healthy donors, the distributions of the two groups largely overlapped, suggesting that merely counting is not sufficient to distinguish the cancer patients from healthy donors (Figure 2a). A median of two and five putative neoepitopes were detected in the plasma cfRNA of pancreatic cancer patients and healthy donors, respectively, in the GSE136651 dataset, which is sequenced at only 1/10th the coverage as compared to the GSE174302 dataset (Figure S2a).

We investigated which of the putative neoepitopes were differentially expressed in the cfRNA of cancer patients compared to the healthy donors. We first excluded generic neoepitopes that were highly expressed in cfRNA of cancer patients as well as healthy donors (Supplementary Table S2). Reassuringly, the generic neoepitopes mostly consisted of peptides derived from mutations in the mitochondrial genes (Supplementary Table S2). Then putative neoepitopes with >=2-fold differential expression in the cancer patients vs. healthy donors were retrieved (see Methods). Among the neoepitopes that were differentially expressed in HCC, nine (six shown) came from MUC12, ‘mucin 12 cell surface associated’ protein, seven (six shown) came from HRNR, ‘hornerin’ protein and seven (five shown) came from OR4C5, ‘olfactory receptor family 4 subfamily C member 5’ protein (Figure 2c, Supplementary Table S5a). There were only a few putative neoepitopes enriched in the healthy donors, but surprisingly one of them was from TPT1, ‘tumour protein translationally controlled 1’ protein. Similarly, discriminative neoepitopes were detected in cfRNA of other cancer types (Supplementary Figure S2, S3 and Table S5). Among the 250 putative neoepitopes that were differential expressed in patients from all cancer types vs. healthy, only 11 were differentially expressed in all 6 cancer types (Supplementary Table S5g and Figure S4).

### Benchmarking the tools for detecting known neoepitopes

We benchmarked the nullomer approach for neoepitope detection against the two most widely used variant discovery tools, gatk haplotypeCaller (41) and bcftools mpileup (42). In addition, we benchmarked against Lofreq (43), which is especially well suited for low frequency variants. Interestingly, there was little overlap between the putative neoepitopes detected by the four methods, with the nullomer approach having the least overlap with the other three (Supplementary Figure S5a & c). There were only 71 unique putative neoepitopes detected by all four methods in the GSE142987 dataset, and 12 in the GSE136651 dataset (Supplementary Table S6). This is not a surprise as the nullomer approach is much different than the other three, which share most of the components of their pipelines.

Notable differences include the fact that our approach uses stringent filters based on base qualities, alignment score and mapped length. Our approach is sensitive to read orientation, it ignores the antisense reads, and maps the reads to the CCDS using bowtie2 (39). All other tools shown here for the comparison use STAR (45), which does a transcriptome-aware genome alignment. To test the difference in the mapping strategies, we used STAR to map the nullomer containing reads and then converted the genome coordinates to CCDS coordinates to estimate the read coverage of neoepitopes using nullomers. Using STAR mapping increased the common neoepitopes that were detected by all tools by 24% in GSE142987 and 120% in GSE136651 (Supplementary Figure S5b & S5d). 51-53% of the neoepitope-donor pairs that were detected using bowtie were also detected using STAR.

Another notable difference is that most tools filter out reads mapped with a low MAPQ score, which is highly biased against short reads. As cfRNA is mostly fragmented, the short reads are especially important. Hence, we devised a customized alignment-score based filter, which adjusts to the variations in read length. Furthermore, we also search for known neoepitopes arising from two missense variants in the same reads. As expected, the nullomer based approach is much faster than any of the other approaches (Supplementary Figure S6).

### Gene fusions in cfRNA

Next, we used the nullomer approach to detect putative gene fusions in the plasma cfRNA of cancer patients and healthy donors. Similar to before, we restrict our search to the gene fusions published and curated in the ChimerDB database (38). The total number of putative gene fusions detected was small, with no more than four events in any of the patients (Figure 3). The known clinically relevant gene fusions discovered in the cfRNA data consisted of EML4-ALK fusion in one HCC patient and ETV6 fusions in one stomach cancer, nine HCC and four colorectal cancer patients (14, 55–57). The low numbers were not surprising as it has been shown that gene fusions drive cancer progression in only 16.5% of cases (58). Overall, gene fusions were more common in the cfRNA of cancer patients as compared to healthy individuals, consistent with previous studies (58). NAB2 and STAT6 gene fusion in chr12 (between 57486364: + strand and 57490916, - strand) was detected in healthy donors and HCC patients from two of the three merged datasets. Interestingly, seven of the eight gene fusion events detected in the healthy individuals involved the fusion of NAB2 and STAT6 (Figure 3). The third common fusion was YPEL5-PPP1CB, which was detected in nine different cancer patients (Supplementary Table S7) and has also been described as a ‘recurrent reciprocal RNA chimera’ (Velusamy et al., 2013).

**Figure 3.**
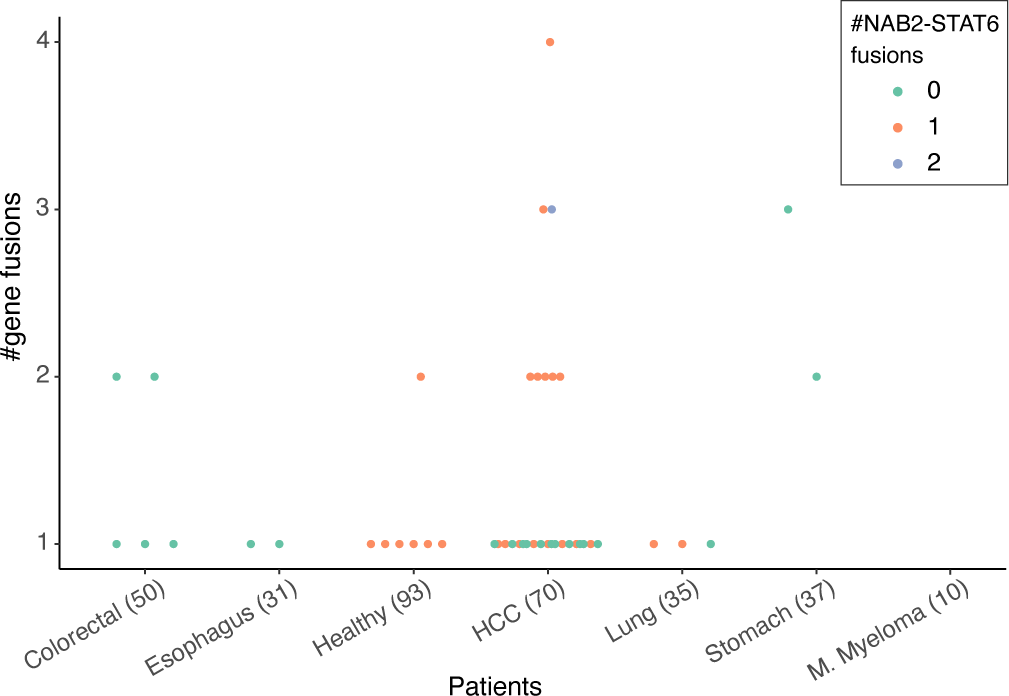
Number of putative gene fusions per patient grouped by healthy donors and cancer type. Patients with NAB2-STAT6 gene fusions are coloured as per legend.

### Transposable elements in cfRNA

The plasma cfRNA from both healthy donors and cancer patients was found to be rich in TEs. Among the 687 TE classes quantified by SalmonTE (48), 279 were found to be differentially expressed between healthy donors and cancer patients (Supplementary Table S8). The selected TEs, shown in Figure 4, are among the most differentially expressed in multiple cancer types. The expression of most TE classes was significantly higher in HCC patients compared to healthy individuals (Supplementary Table S8a). Interestingly, 35, 45, and 5 Alu TEs were found to be differentially expressed in the cfRNA of pancreatic cancer, HCC, and colorectal cancer patients, respectively (Supplementary Figure S7 and Table S8). Similarly, 11, 13, 60, and 19 long terminal repeat (LTR) TEs were differentially expressed in cfRNA of stomach cancer, lung cancer, HCC, and colorectal cancer patients, respectively (Supplementary Table S8a). Finally, 26, 11, and 1 HERV TEs were differentially expressed in HCC and colorectal cancer, and stomach cancer patients, respectively (Supplementary Table S8a). Expression of some TE classes, such as HERVIP10F, were significantly lower in colorectal cancer and stomach cancer patients as compared to the healthy donors (Figure 4; Supplementary data: Table S8a).

**Figure 4.**
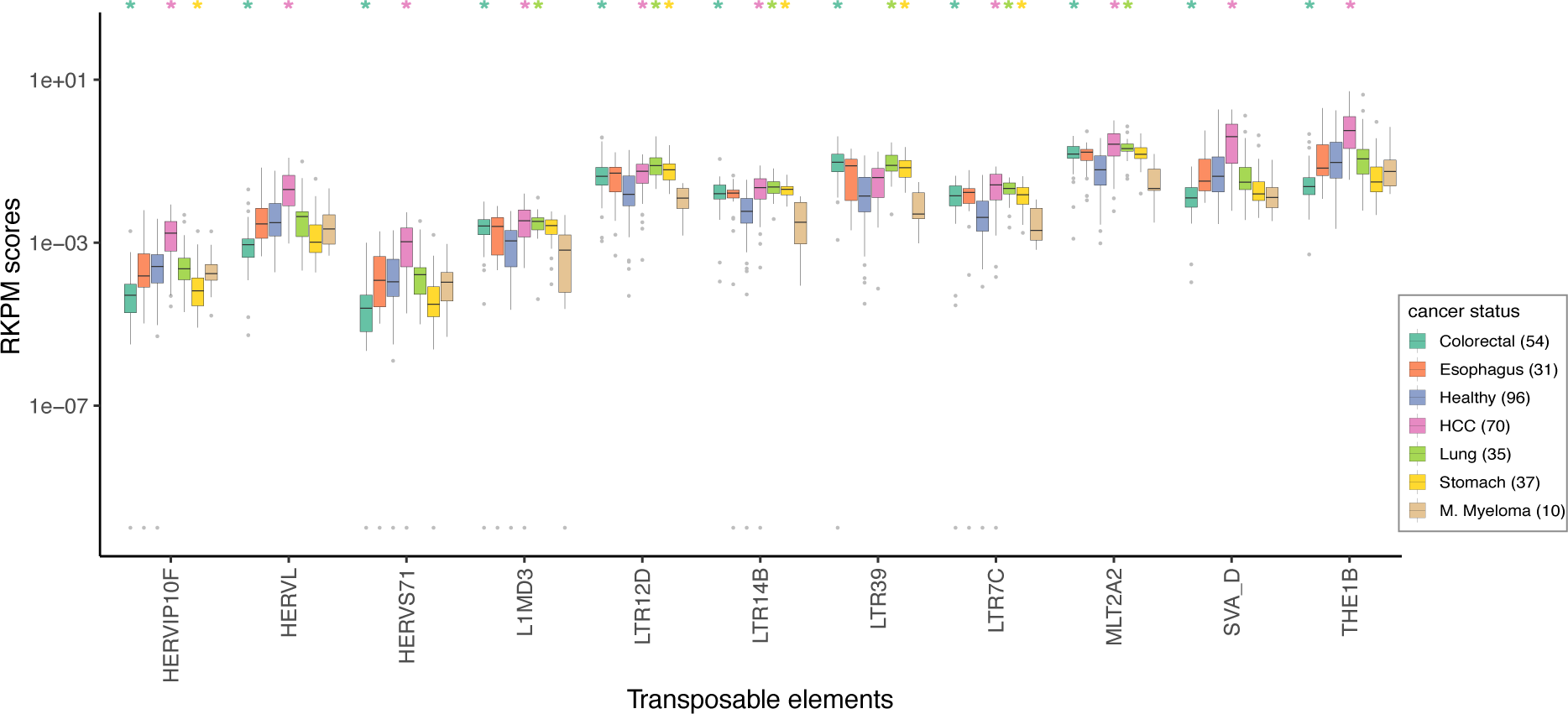
A box and whisker plot showing the top TEs that are differentially expressed (Wilcoxon test corrected p-value <1e-08) in the cfRNA of healthy donors vs. patients of any two cancer types. Only 4 of the 10 such LTR TEs have been shown (see Supplementary Table S8). The cancer-types with significantly different distribution of the TE expression are marked on top with the colour matched asterisk. Outliers with RKPM score <=1e-10 were assumed to have RKPM score =1e-10. The horizontal line corresponds to the median, two hinges correspond to the 25th and 75th percentiles, and two whiskers correspond to the largest and smallest value no further than 1.5 times the distance between 25th and 75th percentiles.

### circRNAs in cfRNA

CIRIquant was used for de-novo detection of circRNAs in the cfRNA (15). The number of circRNAs detected in the cfRNA of HCC patients varied from 1 to ∼1,000 and the circRNAs detected in healthy donors followed a similar distribution (Figure 5a). The number of circRNAs detected in the cfRNA of Colorectal cancer patients were significantly higher than healthy donors (Figure 5a), and the first two PCs were able to partially segregate the cluster of colorectal cancer patients from the healthy donors (Figure 5b). Even while using moderate cutoffs for p-value and recurrence in groups of donors, only a few circRNAs were found to be differentially expressed in cancer patients against the healthy controls (Supplementary Table S9). Differentially expressed circRNAs were only found in colorectal, stomach and lung cancer, and ‘ZCCHC7’ was the only one such circRNA that was found in HCC (Supplementary Table S9).

**Figure 5.**
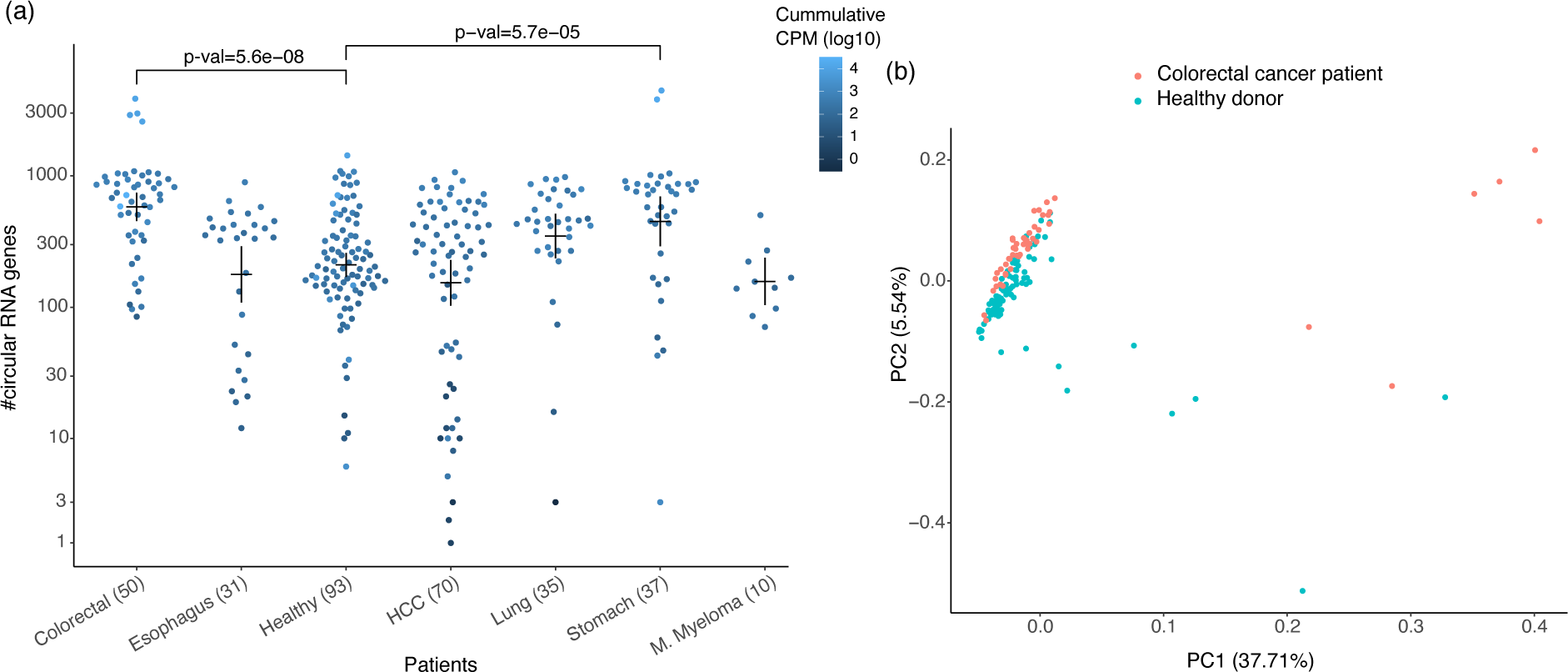
(a) Number of predicted circRNAs in the cfRNA of patients grouped by cancer type and healthy. Each dot is coloured by cumulative CPM of all the circRNAs in the respective cfRNA sample. P-values of two cancer types with most significant differences in distribution of circRNAs in healthy donors vs. patients are shown. P-values were calculated using the Wilcoxon test with Bonferroni correction. Means and error-bar are shown for each group. (b) First two components of a PCA using log of CPM scores of all circRNAs detected in the colorectal cancer patients and healthy donors.

### Clustering and supervised learning

To demonstrate that it is possible to discriminate between healthy and cancer samples based on the biomarkers discovered in cfRNA, we developed classifiers using expression of putative neoepitopes TEs and circRNAs as features. Gene fusions were not used for classification due to low recurrency among the samples used in this study, but they can easily be incorporated into our framework. The classifiers trained using all three modalities performed the best. Using five-fold cross validation, random forest classifiers reached a balanced accuracy of 0.81 and 0.84 in HCC and colorectal cancer, respectively (Table 1; Supplementary Tables S14). The classifiers were able to identify cancer patients with high sensitivity and specificity. Among 96 healthy donors and 66 HCC patients in the merged dataset, a selected TE was expressed in average 112.52 (median 126) cfRNA donors, while a selected neoepitope was only expressed in average 3.24 (median 1) cfRNA donors. Thus, the classifier must be able to handle this difference in recurrence of TE and neoepitope expression. The performance of Ridge and Lasso classifiers suffers while training on a merged feature-set, as the optimal value of regularisation parameter lambda differs for the TEs and neoepitopes. The random forest classifier performed overall better than SVM, Lasso and Ridge regression (Supplementary Table S9-14). We also observed that samples with little or no expression of the selected features were the ones that were prone to wrong classification.

**Table 1:** Random forest classifier performance averaged from 10 iterations of 5-fold cross-validation. As the performance metrics were estimated for three different sets of features, samples with no expressed features were included.

| Cancer Type | Features | F1 | Balanced accuracy | Specificity | Sensitivity |
| --- | --- | --- | --- | --- | --- |
| HCC | neoepitopes | 0.80 | 0.75 | 0.63 | 0.86 |
| HCC | TEs | 0.85 | 0.81 | 0.74 | 0.87 |
| HCC | circRNA | 0.72 | 0.65 | 0.56 | 0.74 |
| HCC | neoepitopes+TEs | 0.84 | 0.81 | 0.80 | 0.83 |
| HCC | neoepitopes+TEs+circRNA | 0.84 | 0.81 | 0.79 | 0.84 |
| Colorectal | neoepitopes | 0.82 | 0.76 | 0.70 | 0.83 |
| Colorectal | TEs | 0.85 | 0.78 | 0.71 | 0.86 |
| Colorectal | circRNA | 0.83 | 0.75 | 0.63 | 0.86 |
| Colorectal | neoepitopes+TEs | 0.87 | 0.82 | 0.76 | 0.88 |
| Colorectal | neoepitopes+TEs+circRNA | 0.88 | 0.84 | 0.80 | 0.87 |

The expression level of human and microbe derived transcripts in plasma cfRNA has been shown to achieve an AUROC > 90% for distinguishing HCC and colorectal cancer patients from the healthy controls (8). In another study, expression of 3 ncRNAs (SNORD3B-1, circ-0080695, and miR-122) obtained the highest average AUROC of 89.4% (6). In comparison, our classifier trained on putative neoepitopes, TEs, and circRNAs in plasma cfRNA achieved an average AUROC of 87% for HCC and 92% for colorectal cancer (Figure 6, Supplementary Tables S14e & j).

**Figure 6.**
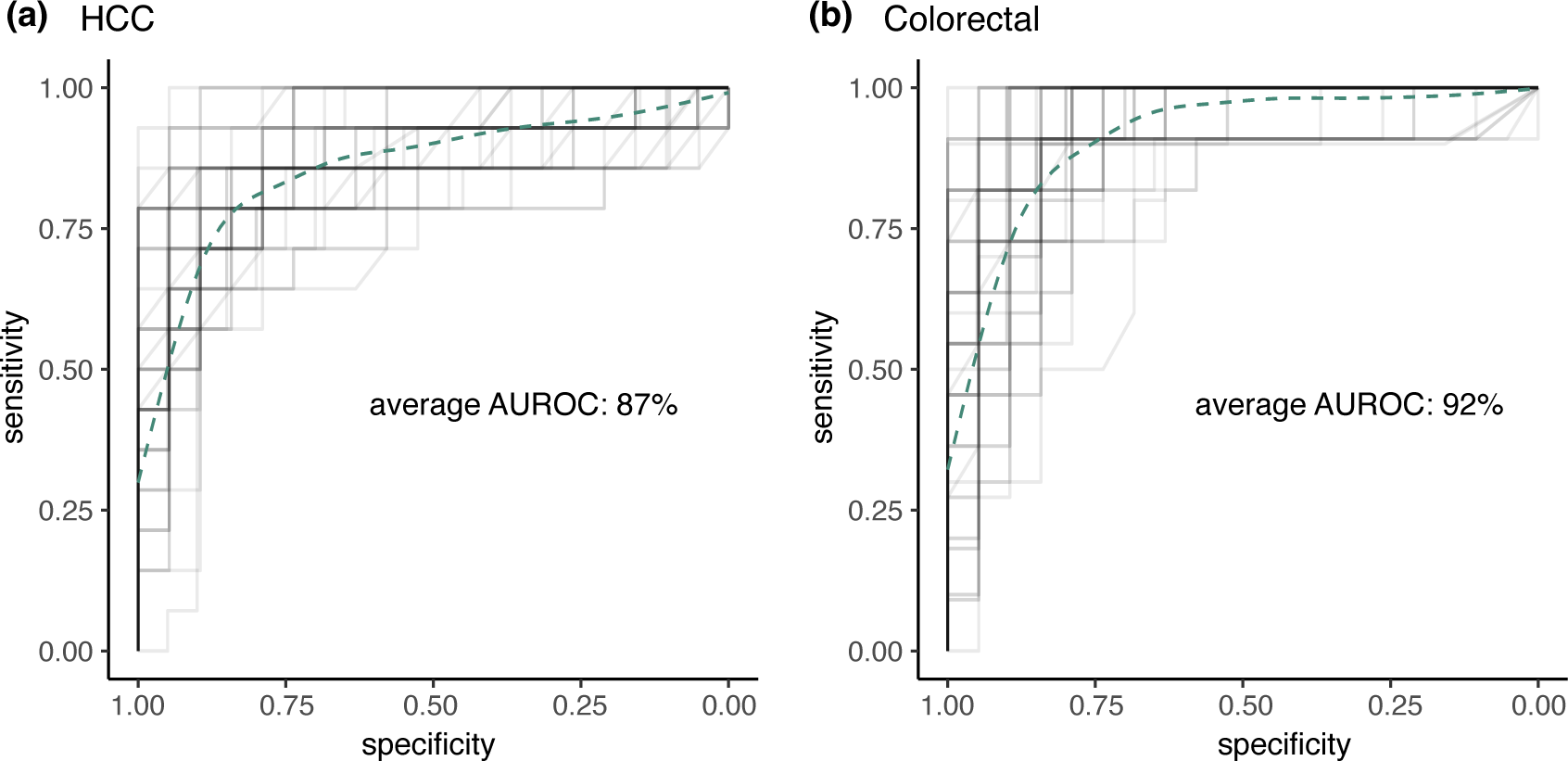
Overlapping ROCs of all 5 folds of the 10 iterations of 5-fold cross validation of the random forest classifier while classifying (a) HCC patients, and (b) colorectal cancer patients among the healthy donors. The classifier was trained using expression of neoepitopes, circRNAs and TEs in the cfRNA samples. Each ROC line was plotted with alpha=0.1 and the smooth spline has been generated by fitting a generalized additive model to all the ROCs.

To test the classifier performance across batches, a random forest classifier was trained for HCC detection using donors from two of the datasets at a time and tested on the remaining one dataset. The merged set of features was used with the same selection strategy as above. The best performing classifier was the one trained on the features detected in cfRNA of 35 HCC patients, and 66 healthy donors in the GSE182824 and GSE174302 datasets. It achieved a balanced accuracy of 0.68 when tested on the 35 HCC patients, and 30 healthy donors in the GSE142987 dataset (Supplementary Table S15). 20 out of 103 putative neoepitopes that were selected to train the above classifier were not expressed in any of the samples in the test-set. Overall, the results suggest that batch specific noise is present in the cfRNA, however, features that are recurrent in a disease-specific manner are also present.

## DISCUSSION

Cancer causes systemic changes, some of which can be detected in the cfRNA from body fluids. In this study we have characterised some of the biomarkers that can be derived from cfRNA and used to detect cancer. We used nullomers to identify putative neoepitopes, and we demonstrated that the composition of putative neoepitopes can be used to distinguish cancer patients from the healthy controls. Nullomers were also utilised to identify putative gene fusions, which are hard to detect in RNAseq data (59), and cannot be easily distinguished from other chimeric RNA (60). Although the number of detected gene fusions was small, they were mostly found in cancer patients.

We showed that the TEs and circRNAs in the cfRNA are also rich in cancer-specific biomarkers. We combined these two independent modalities with the neoepitopes, to obtain a classifier with a balanced accuracy of 0.84 for colorectal cancer, and 0.81 for HCC. Among these three modalities, the cancer classifiers trained using only TE expression performed best, and the classifiers trained using only circRNAs performed the worst. Currently available tools for circRNA prediction are known for low sensitivity and require RNAse P treated samples to improve the accuracy (61).

We introduce a nullomer-based approach for rapid detection of known neoepitopes and gene fusions directly in the cfRNA data. The advantage of using the nullomer-based approach is that it does not require any genotype information, is inclusive of fragmented short reads, uses stringent criteria to filter variants arising due to technical artefacts and filters germline variants. In summary, the nullomer approach is fast, has minimal dependencies, and finds neoepitopes that existing tools tend to miss.

Some promising patterns were observed but substantially more cfRNA data is needed from both healthy donors and patients from multiple cancer types and stages to be able to more confidently assess the accuracy of our method. Compared to the classifiers developed by the authors of the dataset that we analysed, we found that our method has similar performance. A key difference is that our framework is generally applicable across cancer types. Moreover, our method provides additional, clinically relevant information in the form of putative neoepitopes. However, the cells that produce the discriminative neoepitopes, and their ability to activate immune cells against tumours must be verified experimentally. The growing repertoire of experimental validated neoepitopes and gene fusions may also improve the predictions of this method. An important validation experiment to follow up on this study is to profile the tumour as well, ideally with mass-spectrometry, to validate that the predicted neoepitopes are indeed displayed on the malignant cells. To the best of our knowledge no such dataset exists in the public domain today, so both blood and tumour samples would have to be first collected and profiled.

Another advantage of our method is that it is independent of other modalities, and as such it can be combined with approaches that consider DNA or protein biomarkers in a liquid biopsy to detect cancer. Together with RNA from peripheral immune cells and RNA from circulating intact tumour cells cfRNA can provide additional biomarkers that could be tracked during cancer diagnosis and progression. It has become clear that raw tumor mutation burden is a poor predictor of cancer diagnosis, tumor foreignness and response to immunotherapy however, biologically relevant subsets of the tumor mutational burden, such as neoepitope producing mutations might provide a more reliable predictive metrics (62). Overall, detecting relevant biomarkers in cfRNA and tracking them over a period could be beneficial in disease detection and prognosis.

## Data Availability

All data produced in the present study are available upon reasonable request to the authors

## DATA AVAILABILITY

The FastNeo package is available at https://github.com/yashumayank/FastNeo and https://zenodo.org/records/11521368. The package also contains the benchmark pipelines to detect IEDB and TSNAdb neoepitopes using HaplotypeCaller, bcftools and Lofreq, and to run FastNeo with STAR instead of Bowtie2.

## AUTHOR CONTRIBUTIONS

MM wrote the code and analysed the data. MH developed the concept and supervised the research. MM and MH wrote the manuscript together.

